# Artificial Intelligence based personalized diet: A pilot clinical study for IBS

**DOI:** 10.1101/2021.02.23.21251434

**Authors:** Tarkan Karakan, Aycan Gundogdu, Hakan Alagözlü, Nergis Ekmen, Seckin Ozgul, Mehmet Hora, Damla Beyazgul, O. Ufuk Nalbantoglu

## Abstract

**Background and aims:** Certain diets often used to manage functional gastrointestinal symptoms in patients with irritable bowel syndrome (IBS). Personalized diet-induced microbiome modulation is being preferred method for symptom improvement in IBS. Although personalized nutritional therapies targeting gut microbiota using artificial intelligence (AI) promises a great potential, this approach has not been studied in patients with IBS. Therefore, in this study we investigated the efficacy of AI-based personalized microbiome diet in patients with IBS-Mix (M).

**Methods:** This study was designed as a pilot, open-labelled study. We enrolled consecutive IBS-M patients (n=25, 19 females, 46.06 ± 13.11 years) according to Rome IV criteria. Fecal samples were obtained from all patients twice (pre- and post-intervention) and high-througput 16S rRNA sequencing was performed. Patients were divided into two groups based on age, gender and microbiome matched. Six weeks of AI-based microbiome diet (n=14) for group 1 and standard IBS diet (Control group, n=11) for group 2 were followed. AI-based diet was designed based on optimizing a personalized nutritional strategy by an algorithm regarding individual gut microbiome features. An algorithm assessing an IBS index score using microbiome composition attempted to design the optimized diets based on modulating microbiome towards the healthy scores. Baseline and post-intervention IBS-SSS (symptom severity scale) scores and fecal microbiome analyses were compared.

**Results:** The IBS-SSS evaluation for both pre- and post-intervention exhibited significant improvement (p<0.02 and p<0.001 for the control and intervention groups, respectively). While the IBS-SSS evaluation changed to moderate from severe in 82% (14 out of 17) of the intervention group, no such change was observed in the control group. After 6-weeks of intervention, a major shift in microbiota profiles in terms of alfa- or beta-diversity was not observed in both groups. A trend of decrease in Ruminococcaceae family for the intervention group was observed (p=0.17). A statistically significant increase in Faecalibacterium genus was observed in the intervention group (p = 0.04). Bacteroides and putatively probiotic genus Propionibacterium were increased in the intervention group, however Prevotella was increased in the control group. The change (delta) values in IBS-SSS scores (before-after) intervention and control groups are significantly higher in the intervention group.

**Conclusion:** AI-based personalized microbiome modulation through diet significantly improves IBS-related symptoms in patients with IBS-M. Further large scale, randomized placebo-controlled trials with long-term follow-up (durability) are needed.

## 1 Introduction

Irritable bowel syndrome (IBS) is a chronic functional gastrointestinal disorder with negative impact on quality of life and healthcare sources [1]. The exact causes of IBS remain largely unknown. These factors are multifactorial and varied among patients. The pathophysiology of IBS is complex, but recent evidence suggests that the gut microbiome may play an important role in the development, progression and severity of these symptoms [2]. The advent of next-generation sequencing has increased investigations to identify changes in the gut microbiome related to IBS. Some investigators reported increased levels of fecal Streptococcus [3], and Proteobacteria levels in the gut mucosa [4]. IBS severity was also associated with lower alpha-diversity [5]. A recent systematic review of 24 studies performed prior to 2018 have found that while there was some overlap, none of the studies reported the same differences in gut microbiota [6, 7]. This inconsistency can be as a result of unique microbiome composition for each patient and for each disease-state. In other words, discovering disease biomarkers of IBS might be challenging due to diverse and heterogenous microbiome compositions across populations. The second reason for this inconsistency might be due to the interpretation of data in gut microbiome studies is complicated by the dynamic alterations of microbiome over time. For this reason, a snapshot of observations from cross-sectional studies lacks the temporal resolution and do not reflect clinical features in IBS. Diet is an interventional approach with increasing popularity for the treatment of IBS. There are certain evidence-based diets used for IBS-symptom relief. The most popular and studied diet is the FODMAP diet [8]. Although FODMAP diet induces rapid symptom-relief (especially for bloating/distension), it has detrimental effects on gut microbiota (lowering microbiome diversity). Actually, the temporary symptom relief by the FODMAP diet is a consequence of decreased gut abundance of bacterial population and it is not a healthy state for the host.

In order to overcome these microbiome-related inconsistencies in clinical studies, we need to personalize microbiota-modifying therapies. This can be performed by specific personalized diets created by machine-learning approaches, which can handle complex gut microbiome data harboring intrinsic correlations.

In this pilot study, we aimed to manipulate the gut microbiota of IBS patients with an individualized diet. The secondary outcome is to measure the therapeutic effect of this diet on disease-specific parameters.

## 2 Materials and methods

### Study cohorts

This study was designed as a pilot, open-labelled study. We enrolled consecutive IBS-M patients (n=25, 19 females, 46.06 ± 13.11 years) according to Rome IV criteria and a healthy control group (n=34) which were used to model IBS classification models. Healthy group consisted of subjects without chronic diseases affecting microbiome and antibiotic/probiotic consumption in the previous 6 week-period. IBS-M patients were excluded if they had severe cardiac, liver, neurological, psychiatric diseases or a gastrointestinal disease other than IBS (eg celiac disease or inflammatory bowel disease). The patients were not allowed or enrolled if they were following a restricted diet for any purpose. Certain medications involving spasmolytics, antidepressants, etc were allowed if these were at stable doses for the previous 4 weeks. Probiotics and antibiotics (including rifaximin) were not allowed for the previous 6 weeks. Paired fecal samples were obtained (pre- and post-intervention) and high-througput 16S rRNA sequencing was performed to reveal the microbiota compositions at the baseline and post-intervention. Patients were divided into two groups based on age and gender. Moreover, baseline microbiota compositions were clustered to form subpopulations, and each treatment group were populated to represent similar subpopulation diversity. Six weeks of personalized microbiome diet (n=14) for group 1 and standard IBS diet (Control group, n=11) for group 2 were followed.

### Fecal sampling and 16S ribosomal RNA gene sequencing

Fecal samples were collected using BBL culture swabs (Becton, Dickinson and Company, Sparks, MD) and transported to the laboratory in DNA/RNA shield buffer medium. DNA was extracted directly from the stool samples using a Qiagen Power Soil DNA Extraction Kit (Qiagen, Hilden, Germany). The final concentrations of extracted DNA were measured using a NanoDrop (Shimazu). dsDNA quantification was done using the Qubit dsDNA HS Assay Kit and a Qubit 2.0 Fluorimeter (Thermo Fisher Scientific, Waltham, MaA USA) and then they were stored at 20°C for further analysis.

The sequencing of 16S rRNA was performed according to the protocol of the manufacturer (16S Metagenomic Sequencing Library Preparation Preparing 16S Ribosomal RNA Gene Amplicons for the Illumina MiSeq System) using Illumina MiSeq (Illumina, San Diego, CA USA) system. In brief, 2-step PCR amplification were used to construct sequencing library. The 1st step of PCR is to amplify the V4 hypervariable region. The full length of the primers was: 515F, forward 5’ GTGCCAGCMGCCGCGGTAA3’ and 806R, reverse ‘GGACTACHVGGGTWTCTAAT3’ [9]. PCR amplification was performed using a 25L reaction volume that contained 12.5L of 2X KAPA HiFi HotStartReadyMix (KAPA Biosystems, Wilmington, MA USA), 0.2M each of forward and reverse primer, and 100ng of the DNA template. The reaction process was executed by raising the solution temperature to 95°C for 3min, then performing 25 cycles of 98°C for 20sec, 55°C for 30sec, and 72°C for 30sec, ending with the temperature held at 72°C for 5min. Amplicons were purified using the AMPure XP PCR Purification Kit (Beckman Coulter Life Sciences, Indianapolis, IN USA). The second step of PCR is to add the index adaptors using a 10-cycle PCR program. The PCR step adds the index 1 (i7), index 2 (i5), sequencing, and common adapters (P5 and P7) and PCR amplification was performed on a 25L reaction volume containing 12.5L of 2X KAPA HiFi HotStartReadyMix (KAPA Biosystems, Wilmington, MA USA), 0.2M of each index adaptor (i5 and i7), and 2.5L of the first-PCR final product. The reaction process was executed by raising the solution temperature to 95°C for 3min, then performing 10 cycles of 98°C for 20sec, 55°C for 30sec, and 72°C for 30sec, ending with a 72°C hold for 5min. Amplicons were purified using the AMPure XP PCR Purification Kit (Beckman Coulter Life Sciences, Indianapolis, IN USA).

All amplified products were then checked with 2% agarose gel electrophoresis then amplicons were purified using the AMPure XP PCR Purification Kit (Beckman Coulter Genomics, Danvers, MA, USA) and quantified using the Qubit dsDNA HS Assay Kit and a Qubit 2.0 Fluorimeter (Thermo Fisher Scientific, Waltham, MA USA). Approximately 15% PhiX Control library (v3) (Illumina, San Diego, CA USA) was combined with the final sequencing library. The libraries were processed for cluster generation and sequencing on 250PE MiSeq runs, and generating at least 50.000 reads per sample.

Sequencing data were analyzed using QIIME pipeline [10] after filtering and trimming the reads for PHRED quality score 30 via Trimmomatic tool [11]. Operational taxonomic unites were determined using Uclust method and the units were assigned to taxonomic clades via PyNAST using Green Genes database [12] with open reference procedure. Alpha- and beta-diversity statistics were assesed accordingly by QIIME pipeline scripts. The graph-based visualization of the microbiota profiles was performed using tmap topological data analysis framework () with Bray-Curtis distance metric.

### IBS-index Scoring

The baseline group of IBS-M patients (n=25) and the healthy controls (n=34) were compared in terms of their microbiota compositions. The detected microbiota profiles were used to characterize the disease in a classification setting. Based on Gradient Boosted Trees (GBT) [13] classification algorithm, a stochastic gradient boosting classification model (XGBoost, version 0.90 [14]) was used in dropouts meet multiple additive regression trees (DART) booster with binary logistic regressor. 5-fold cross-validation, with 10 random seeding trials were used to observe the disease classification performance. The logistic regression scores of XGBoost models were used as IBS-index scores. The dataset was utilized to train the final IBS-index model. The hyperparameters of the XGBoost model were optimized using Bayesian optimization tool Optuna [15] in 5-fold-cross validation setting.

### The AI-based personalized nutrition model

Enbiosis personalized nutrition model estimates the optimal micronutrient compositions for a required microbiome modulation. In this study, we computed the microbiome modulation needed for an IBS case, based on the IBS-indices generated by the machine learning models. According to that, the baseline microbiome compositions are perturbed randomly with a small probability p. Perturbed profiles are accepted with a probability proportional to the decrease in the IBS-index as suggested by Metropolis sampling [16]. This Monte-Carlo random walk in the microbiome composition space is expected to meet a low IBS-index microbiome composition nearby the baseline microbiome composition of the patient with a minimal modulation. The personalized nutrition model, then, estimates the optimized nutritional composition needed for this individual, expecting to drive the IBS-index to lower values.

Therefore, an algorithm assessing an IBS index score using microbiome composition attempted to design the optimized diets based on modulating microbiome towards the healthy scores.

## 3 Results

### Gut microbiota communities between IBS patients and Healthy Controls

The gut microbiome genus level abundance profile is shown in figure 1. The gut microbiome profile of the recruited patients and the healthy controls showed significant differences in beta diversity. Based on unweighted UniFrac dissimilarity measurement of microbiota sample pairs, the patient and the healthy control groups showed different community profiles (*p* < 10^−6^, PERMANOVA test with 1,000,000 random permutations). The stratified profiles can be observed in the tmap visualization in Figure 2. Clear subgroupings between the IBS-cases and the healthy controls can be observed from these topological maps. When bacterial taxa are considered individually, the most significant differences between the IBS and healthy control groups are observed in Ruminococcaceae (*p* = 0.014, Mann-Whitney U-test) and Clostridiaceae (*p* = 0.022, Mann-Whitney U-test) families and Ruminococcus (*p* = 0.023, Mann-Whitney U-test) and Faecalibacterium (*p* = 0.0005, Mann-Whitney U-test) genera (figure 3,4).

**Figure 1:**
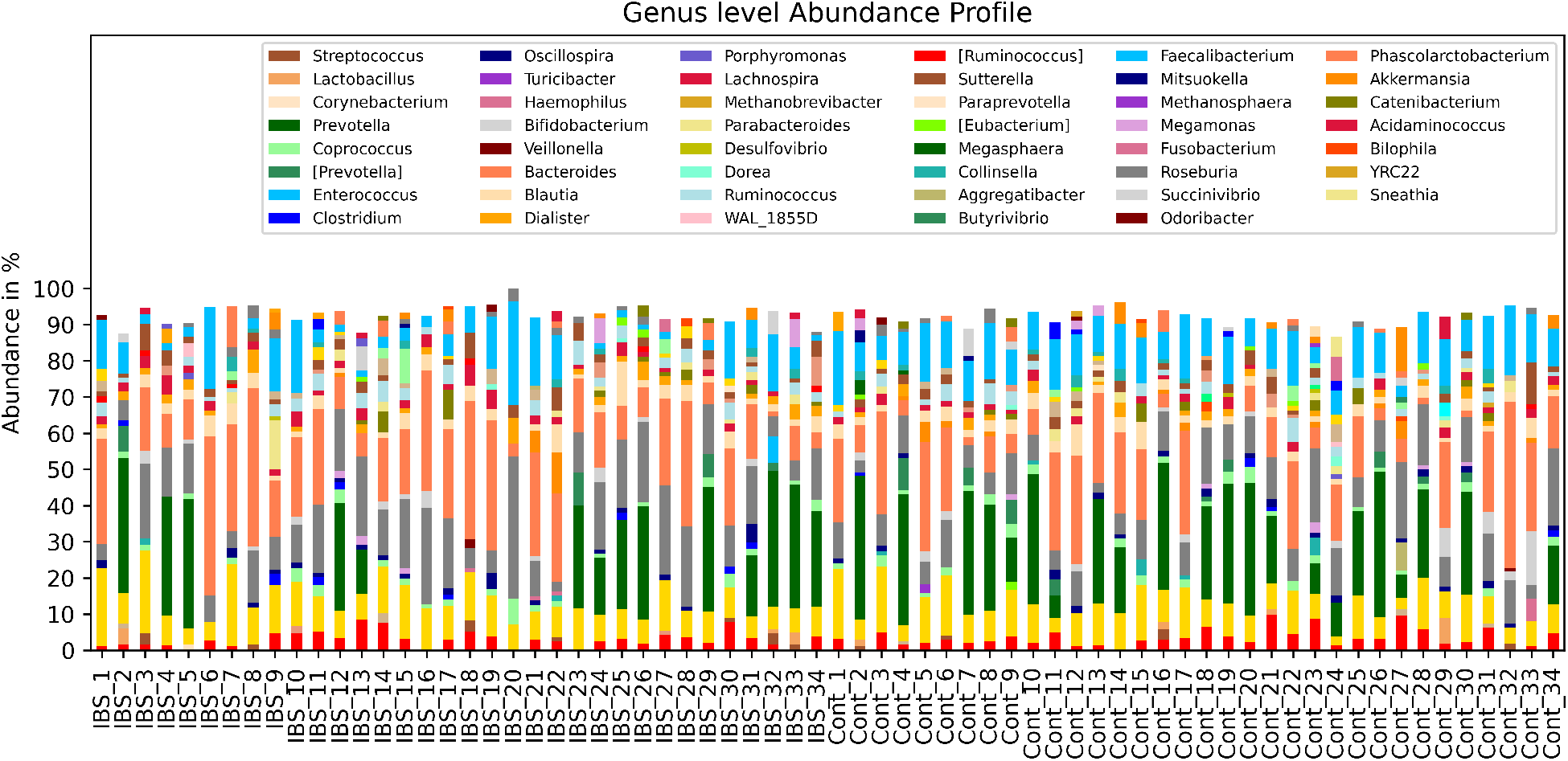
Genus level abundance profiles.

**Figure 2:**
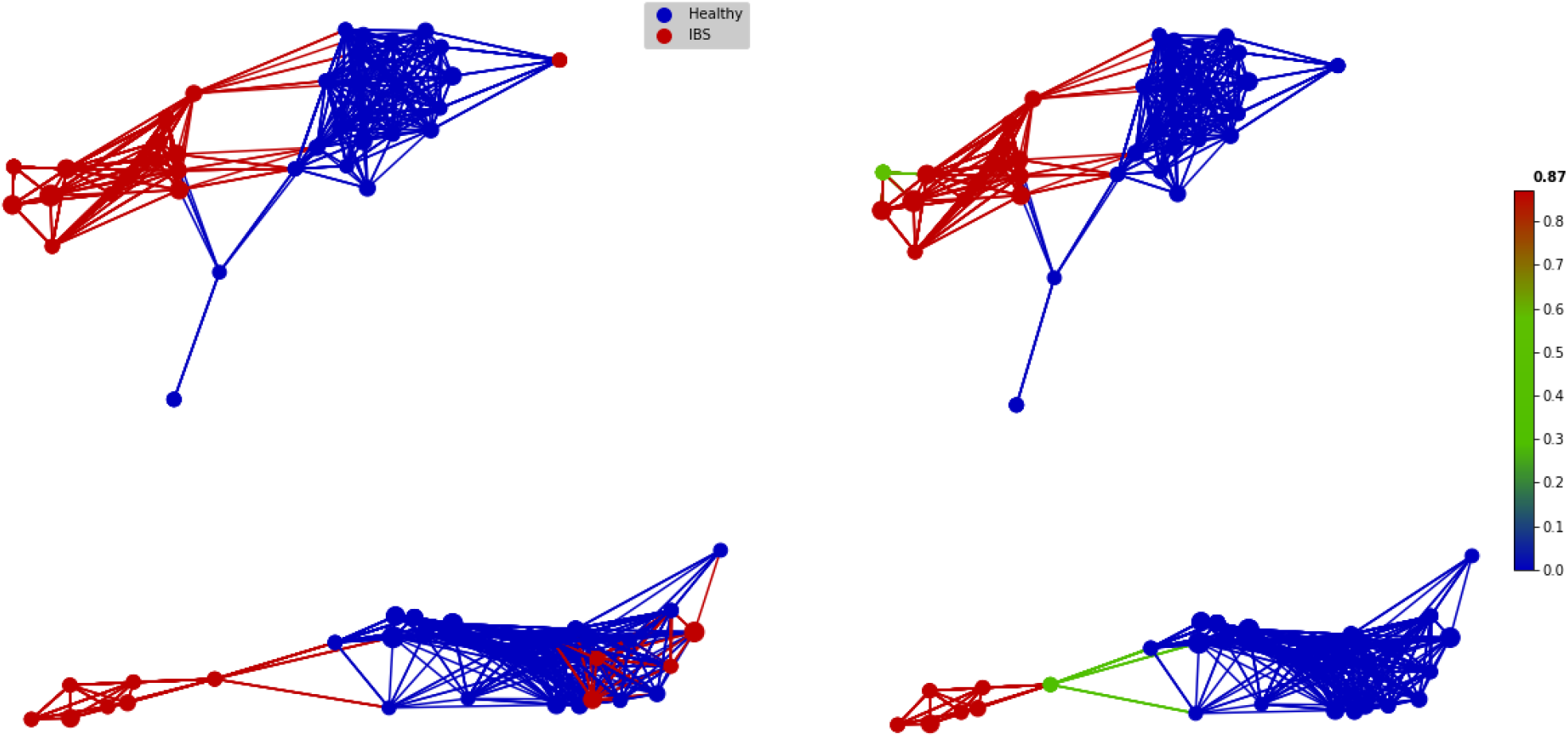
Two-dimensional network visualization of the microbiota profiles using tmap network analysis (constructed by Bray-Curtis metric). Two major enterotypes (up: Bacteroides dominant, down: Preveotella Dominant) nearly form different disease subgroups. Left: network nodes labelled by disease phenotype. Right: SAFE enrichment analysis of the disease scores. Blue-to-red indicates lower to higher IBS scoring.

**Figure 3:**
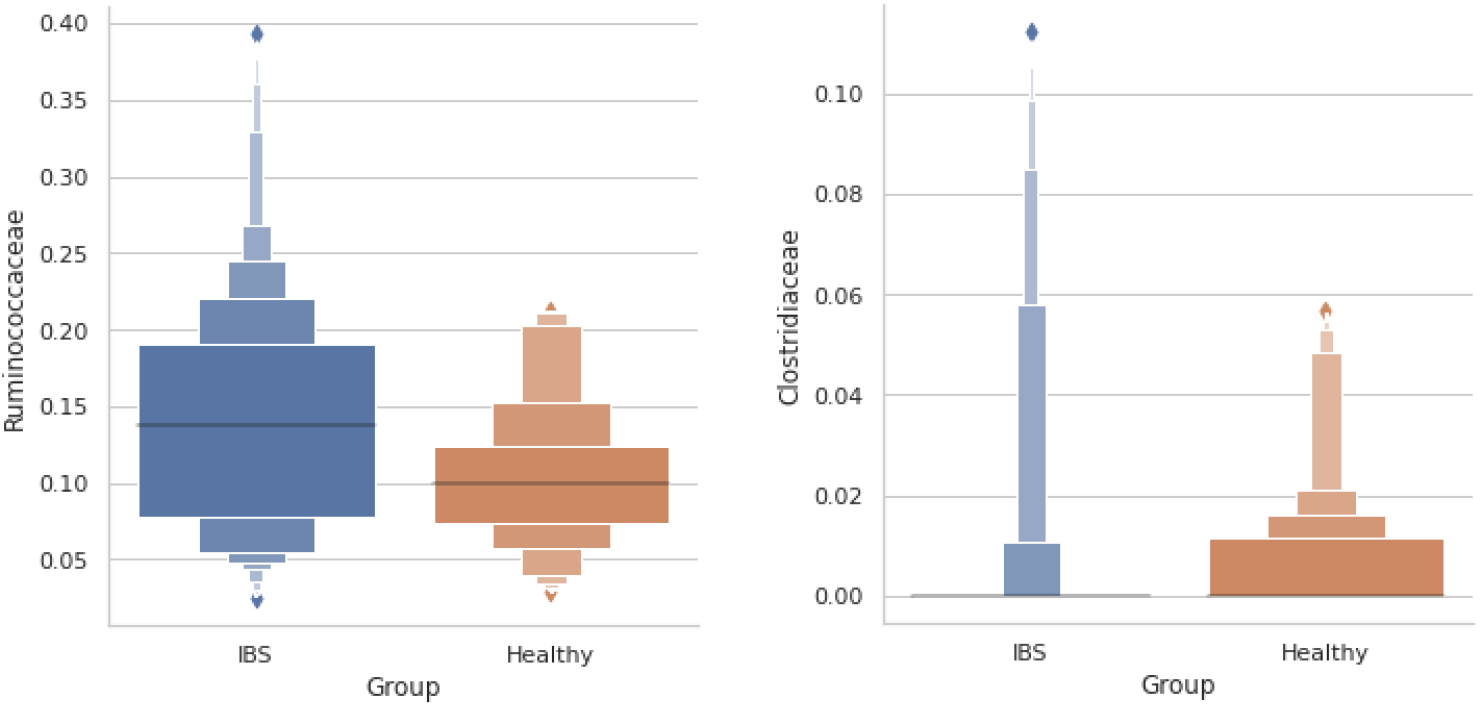
Ruminococcaceae family are observed in higher abundance in IBS group (p-value 0.014, Mann-Whitney u-test), where Clostridiaceae family is decreased in IBS patients (p-value 0.022, Mann-Whitney u-test)

**Figure 4:**
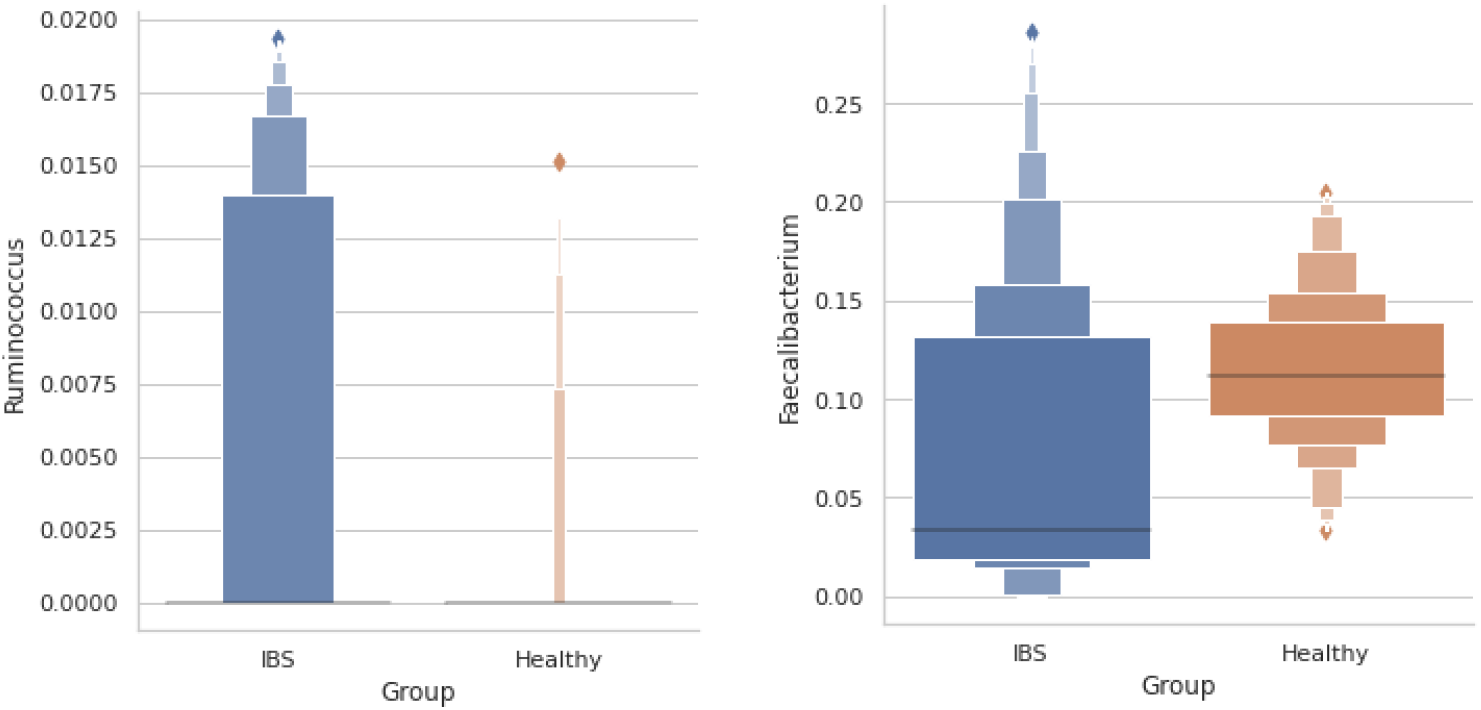
Ruminococcus genus is observed more abundantly in IBS group (p-value 0.023, Mann-Whitney u-test), where Faecalibacterium is observed in significantly lower abundances in IBS patients (p-value 0.0005, Mann-Whitney u-test).

### Disease classification and microbiome derived IBS index scores

A machine-learning (ML) based classifier trained and tested on pre-interventional microbiota profiles exhibited a strong classification performance. Using 5-fold cross validation on the held-out XGBoost classifier models, an average ROC-AUC of 0.964 and an average classification accuracy of 0.91 were determined. The microbiome-derived IBS index scores, which is the inferred disease probability measurements obtained from XGBoost classification models were significantly different (*p* < 10^−5^, Mann-Whitney U-test), which can be seen in Figure 5.

**Figure 5:**
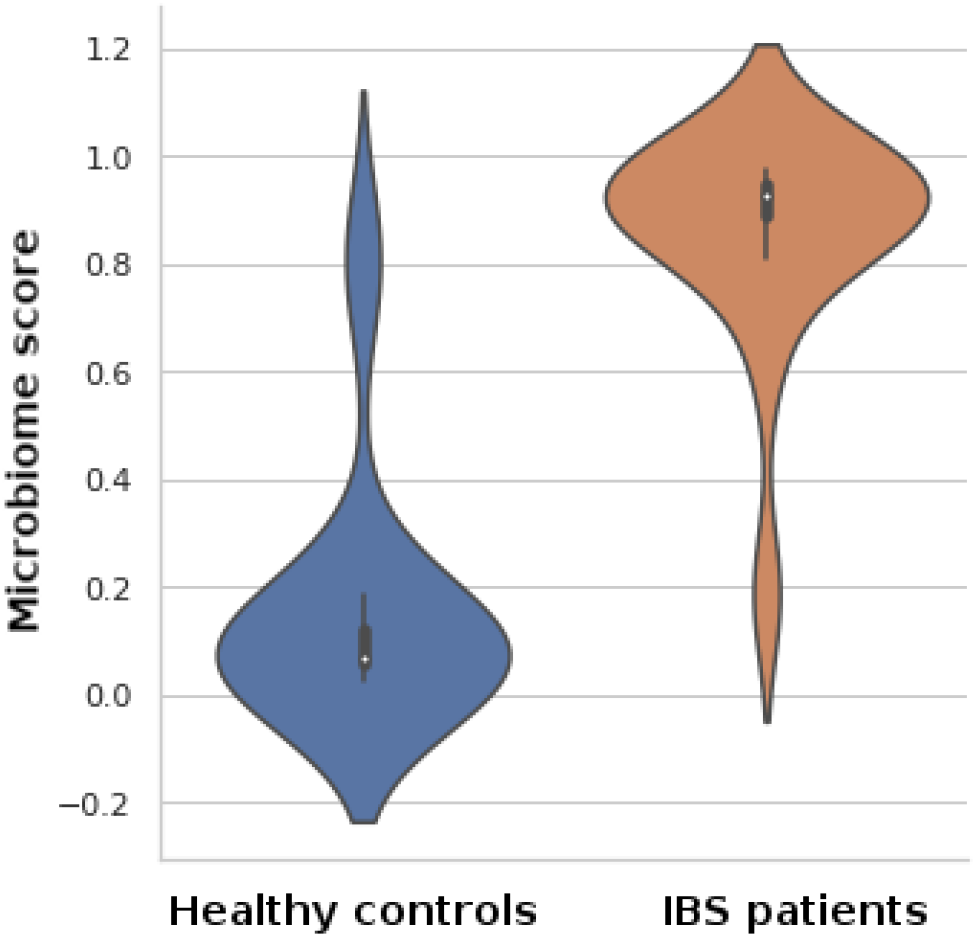
The microbiome scores evaluated for the healthy controls and the IBS patients.

Evaluating the IBS-index scores on the held-out validation cohorts, we observed that the score distributions of the IBS-patients and the healthy controls differ significantly (*p* = 0.001, Mann-Whitney U-test), implying that the machine-learned IBS-index is a strong indicator of the disease.

### Clinical Evaluation of Personalized nutrition vs. control groups

The IBS-SSS evaluation for both pre-intervention and post-intervention conducted for both groups exhibited significant improvement (p<0.02 and p<0.001 for the control and the personalized nutrition interventions respectively). It was observed that the score improvement for the personalized nutrition group was significantly greater than the control group (Table 1, Figure 6).

**Table 1:** IBS-SSS scores (mean ± standard deviation) before and after the interventions.

|  | Pre-intervention | Post-intervention | P-value (paired t-test) |
| --- | --- | --- | --- |
| Personalized nutrition | 357.1 ± 18.2 | 232.5 ± 61.5 | **< 0.001** |
| Control | 363.1 ± 16.7 | 331.8 ± 42.9 | **< 0.02** |

**Figure 6:**
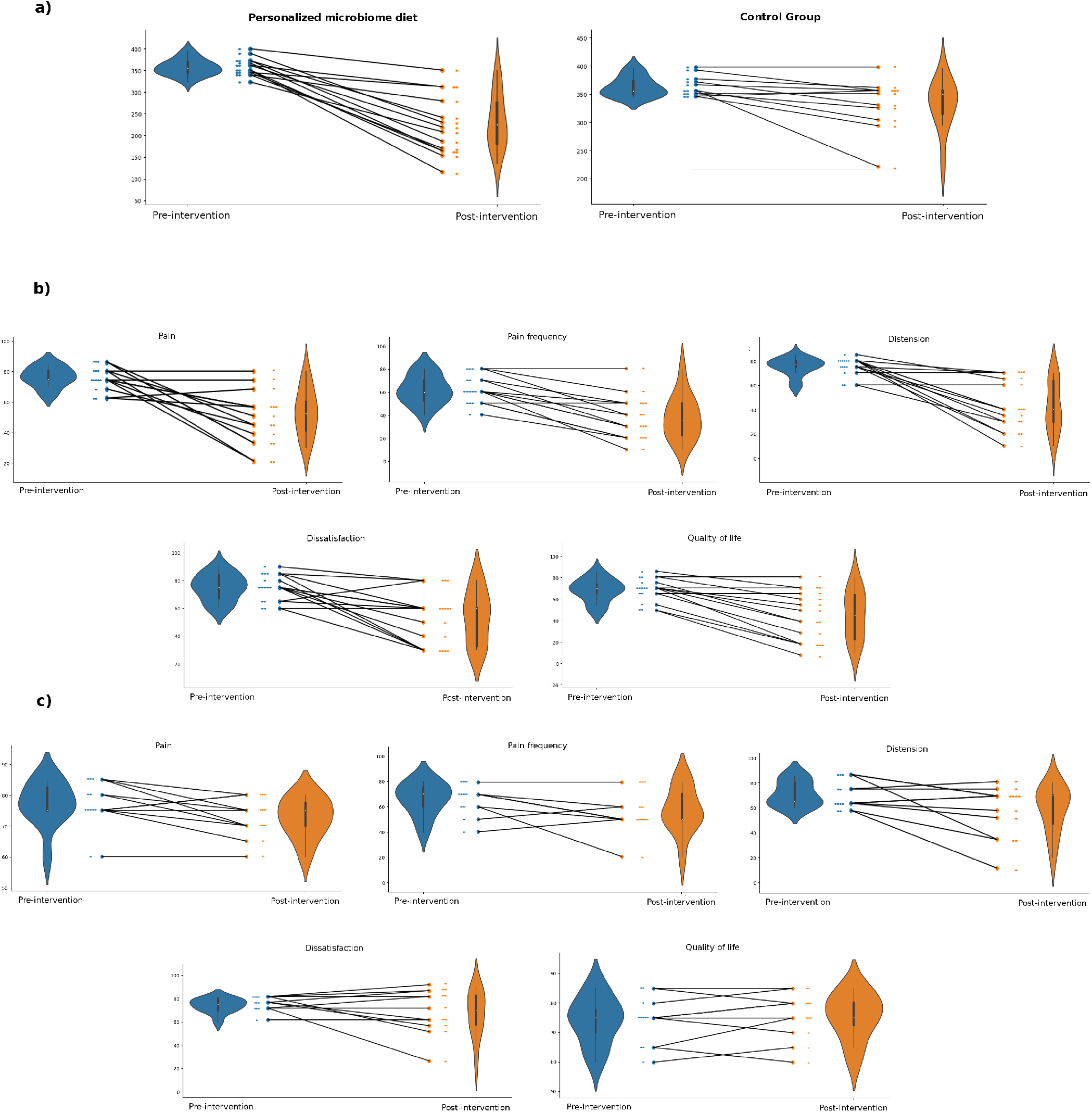
a) IBS-SSS scores for personalized nutrition intervention and IBS-SSS scores for the control intervention. b) IBS-SSS score categories for personalized nutrition pre- and post-intervention. c) IBS-SSS score categories for the control treatment pre- and post-intervention.

Considering each 5 items of IBS-SSS evaluated, the personalized nutrition was observed to be effective on all, whereas abdominal pain frequency, dissatisfaction with bowel habits, and IBS-related quality of life were not changed significantly in the control group (Table 2).

**Table 2:** IBS-SSS score categories (mean ± standard deviation) before and after the interventions.

| Personalized nutrition |  |  |  |
| --- | --- | --- | --- |
|  | Pre-intervention | Post-intervention | P-value (paired t-test) |
| Abdominal pain | 76.4 ± 6.4 | 53.2 ± 15.0 | **< 0.001** |
| Abdominal pain frequency | 62.1 ± 12.0 | 37.9 ± 18.2 | **< 0.001** |
| Distension | 75.4 ± 7.2 | 42.9 ± 19.9 | **< 0.001** |
| Dissatisfaction with bowel habits | 75.0 ± 9.3 | 53.6 ± 18.3 | **< 0.01** |
| IBS-related quality of life | 68.2 ± 10.3 | 45.0 ± 21.7 | **< 0.001** |
| Control |  |  |  |
|  | Pre-intervention | Post-intervention | P-value (paired t-test) |
| Abdominal pain | 77.3 ± 6.7 | 72.7 ± 6.2 | **0.043** |
| Abdominal pain frequency | 66.4 ± 12.3 | 57.3 ± 17.1 | 0.074 |
| Distension | 71.4 ± 9.6 | 59.1 ± 17.7 | **0.041** |
| Dissatisfaction with bowel habits | 74.1 ± 6.0 | 67.3 ± 18.6 | 0.246 |
| IBS-related quality of life | 74.1 ± 7.6 | 75.5 ± 7.5 | 0.391 |

### Post-interventional changes in microbiota profiles

After 6-weeks of intervention, a major shift in microbiota profiles in terms of alfa- or beta-diversity was not observed in both groups. A trend of decrease in Ruminococcaceae family for the personalized nutrition intervention group was observed, however this change was not observed to be statistically significant (*p* = 0.17, paired t-test). A statistically significant increase in Faecalibacterium genus was observed in the personalized nutrition group (*p* = 0.04), where no meaningful change was reported for the control group (*p* = 0.63) (figure 7).

**Figure 7:**
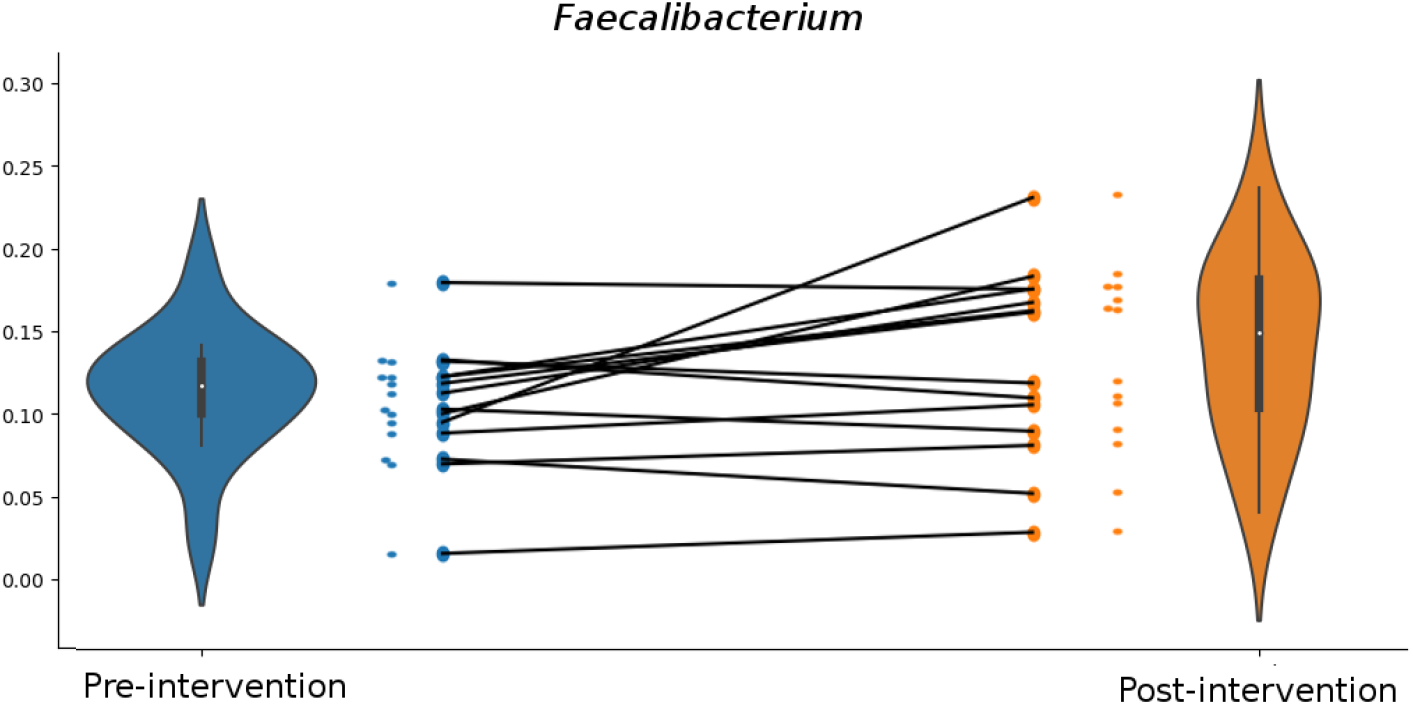
Faecalibacterium relative abundances for the personalized nutrition group pre- and post-intervention.

Both Bacteroides rich and Preveotella rich enterotypes were represented in both personalized nutrition and control intervention groups without significantly different Bacteroides and Prevotella abundances (*p* = 0.34 for Bacteroides and *p* = 0.36 for Prevotella, Mann-Whitney u-test). However, we have observed increase in Bacteroides for the personalized nutrition group (*p >* 0.05) while an increase trend in Prevotella (*p* = 0.057) was noticeable in the control group. Along with that a significant increase for the putatively probiotic genus Propionibacterium (*p* = 0.027) was apparent in the personalized nutrition group where no such increase was observed in the control group.

### The evaluation of microbiome-derived IBS index scores

The microbiome derived IBS index scores both improved towards lower scores in both intervention groups. The improvement in the personalized nutrition group was observed to be greater (Table 3, Figure 8). In order to observe the correlation between microbiome derived IBS scores with the clinical evaluations (i.e. IBS-SSS), we have measured the explained variance of IBS-SSS with respect to microbiome scores (Figure 9). Including the corresponding scores of both intervention groups an *R*^2^ score of 0.652 was found, indicating that the microbiome scores contribute significantly to the explaination of the clinical scores.

**Table 3:** Microbiome scores (mean ± standard deviation) before and after the interventions

|  | Pre-intervention | Post-intervention | P-value (paired t-test) |
| --- | --- | --- | --- |
| Personalized nutrition | $0.89 \pm 0.04$ | $0.62 \pm 0.18$ | **< 0.001** |
| Control | $0.87 \pm 0.05$ | $0.79 \pm 0.11$ | **0.03** |

**Figure 8:**
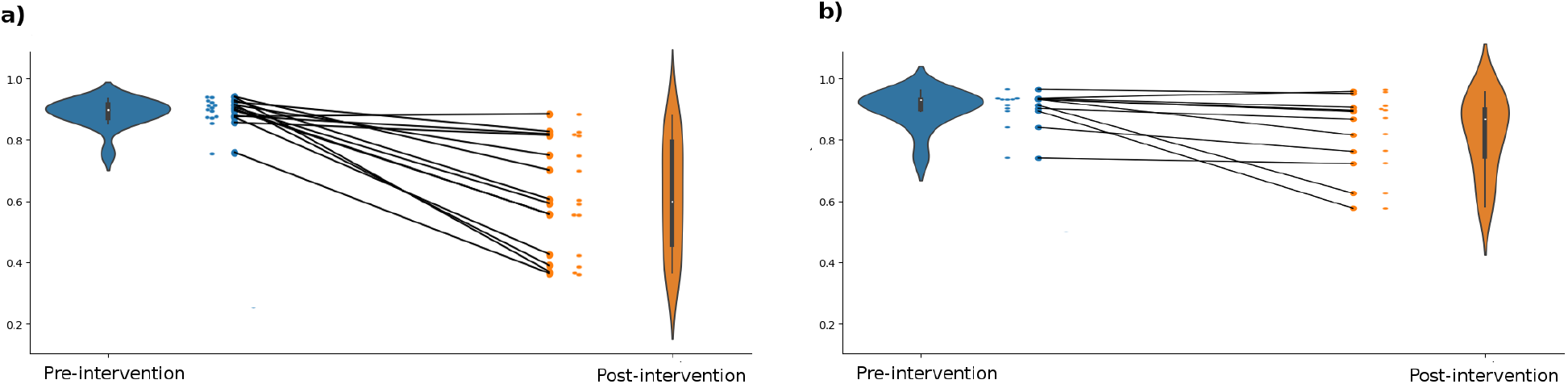
a. Microbiome scores for personalized nutrition intervention, b. Microbiome scores for the control intervention.

**Figure 9:**
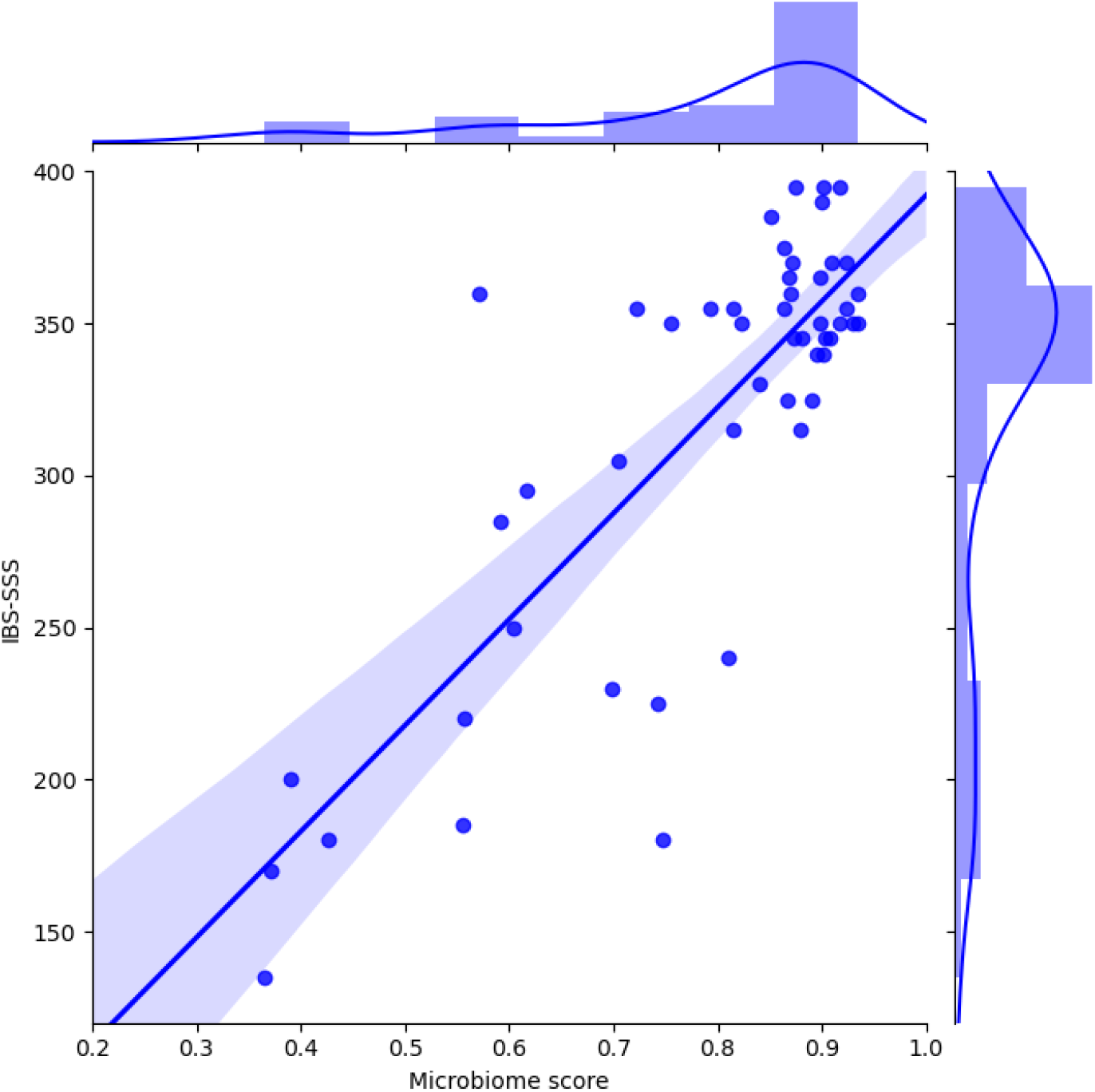
The plot shows the scatter and the marginal histograms of IBS-SSS and microbiome derived IBS scores. The positive correlation is represented by the linear regression line.

## 4 Discussion

Dietary habits constitute a strong driver of interpersonal variance in the gut microbiome composition, and its influence prevails over that of genetics by most estimates [17]. In our study, we investigated the therapeutic effect of the personalized diet on individual gut microbiome and the disease specific symptoms. The majority of IBS patients regard diet as an important trigger for their gastrointestinal symptoms. Based on the subjective correlation with diet and IBS symptoms, there many attempts to design specific diets for the relief of IBS-related complaints. Recent studies indicate that low FODMAP diet relieves some of the IBS symptoms, such as abdominal gas, bloating, distension and even abdominal pain [18]. Elimination of fermentable oligosaccharides, disaccharides, monosaccharides and polyols (FODMAPs) is also recommended by the guidelines [19]. FODMAPs are sugars that ferment in the gut due to inadequate digestion; common ones are lactose, fructose, fructans, and sorbitol. Foods containing FODMAPs include wheat, some fruits and vegetables, corn syrup, and onions. Initial positive study of low FODMAP diet was performed in IBS patients with a positive fructose breath test and without a control group [20]. Another randomized controlled study compared low FODMAP diet with a typical Australian diet and they have found a 30% decrease in IBS-symptom severity [21]. However, subsequent randomized trials failed to detect significant clinical differences between classical IBS diets and low FODMAP diet. All the diets were nearly 50% effective in relieving IBS symptoms and low FODMAP diet is not an exception [22].

Another important but neglected issue about these IBS diets is the diet-related gut microbial changes. In the last decade, there are tremendous amount of studies on gut microbiome in IBS patients [23, 24, 25, 26, 27]. A recent systematic review analyzed 24 studies mostly from Europe and North America. They have found that Bifidobacterium, Faecalibacterium genus are decreased and Lactobacillaceae, Bacteroides, Enterobactericeae family are increased in IBS [28]. In order to overcome the decreased levels of Bifidobacteria, prebiotic or sometimes probiotic supplements might be advised to the IBS patients on low FODMAP diet. While this increases the abundance of Bifidobacteria, it has some deleterious effects gut health in animal studies by disruption of mucosal barrier, increasing mucosal inflammation and visceral hypersensitivity [29]. Rapid colonic fermentation is central to the identified mechanisms that include injury from high luminal concentrations of short-chain fatty acids and low pH, and inflammatory effects of increased endotoxin load and glycation of macromolecules [29].

Today the optimal diet for the treatment of IBS patients is lacking. The ideal diet should be effective on (at least) most of the symptoms in IBS and maintain an eubiotic (or so said healthy) state of gut microbiome. It should be sustainable and personalized.

Our study is the first attempt to reach these therapeutic goals in IBS. We used machine-learning for determining personalized diet to modulate the IBS microbiota to an individually similar “healthy” state. In other words, we tried to formulate a personalized microbiome modulating diet for patients with IBS-M. The gut microbiome genus level IBS and healthy controls showed significant differences in beta diversity. When we look at the bacterial taxa, the most significant differences between the IBS and healthy control groups were observed in Ruminococcaceae and Clostridiaceae families. Ruminococcaceae was increased and Clostridiaceae was decreased in IBS group. In the genera level, Ruminococcus was increased and Faecalibacterium was decreased in IBS group. In a recent systematic review, Ruminococcaceae family and Faecalibacterium genus were not different in IBS vs healthy groups [28]. Although there are inconsistencies between literature and our results, these differences might stem from geographic, cultural and dietary habits of patients.

The IBS-index scores on the held-out validation cohorts were different between IBS-patients and the healthy controls. This implies that the machine-learned IBS-index is a strong indicator of the presence of disease. We detected significant improvement in IBS-SSS values for both pre- and post-intervention periods. The score improvement for the personalized diet group of IBS patients was greater than the control group (Table 1, figure 6). Each of the 5 items of IBS-SSS evaluated, the personalized diet group showed significant improvement on all parameters. However, the control group showed no improvement in abdominal pain frequency, dissatisfaction with bowel habits, and IBS-related quality of life parameters. Böhn et al reported that low FODMAP and standard IBS diet were similar for relieving IBS-symptoms. Abdominal pain frequency and IBS-related quality of life parameters were improved with low FODMAP diet in their study, but the dissatisfaction with bowel habits did not improve [22]. They have noticed a nearly 50% response rate to both diets. This study concluded that low FODMAP diet shows similar clinical benefit with standard IBS diets.

The post-intervention gut microbiome changes were also different between groups. After 6-weeks of intervention, a major shift in microbiota profiles in terms of alfa- or beta-diversity was not observed in both groups. A statistically significant increase in Faecalibacterium genus was observed in the personalized nutrition group (*p* = 0.04), where no meaningful change was reported for the control group (*p* = 0.63). Peter J et al. investigated the role of microbiome in IBS-related psychological distress and found that depression was negatively associated with Lachnospiraceae abundance, the distress, anxiety, depression, and stress perception were associated with higher abundances of Proteobacteria. And the feeling of anxiety was characterized by elevated Bacteroidaceae [30]. In our study, we have observed an increase in Bacteroides for the personalized nutrition group (*p >* 0.05) while an increase trend in Prevotella (*p* = 0.057) was noticeable in the control group. The increase in Bacteroides group might affected the anxiety status of our IBS patients in the intervention group and the improvement in the quality of life scores in IBS-SSS evaluation.

The microbiome derived IBS index scores both improved towards lower scores in both intervention groups. The improvement in the personalized nutrition group was observed to be greater. IBS severity is also correlated with gut microbiome features. Tap J et al. investigated the correlation between gut microbiota signatures and IBS severity. They found that IBS symptom severity to be associated negatively with microbial richness, exhaled CH4, presence of methanogens, and enterotypes enriched with Clostridiales or Prevotella species. This microbiota signature could not be explained by differences in diet or use of medications [31]. In our study, post-interventional analysis showed a increasing trend of Prevotella species (although statistically insignificant) in the control group.

As a result, our study is the first trial in the literature comparing the therapeutic effect of AI-based personalized diet for patients with IBS-M. We had limited clinical and gut microbiome-related benefits after 6-weeks of intervention. Further larger randomized controlled trials are needed to determine the safety, effectiveness and durability of this treatment.

## Data Availability

Data is available upon request.

## Notes

### Competing Interest Statement

The authors have declared no competing interest.

### Clinical Trial

NCT04768387

### Funding Statement

The work was funded by Enbiosis Biotechnology.

### Author Declarations

The approval for the clinical study is cleared by Gazi University Faculty of Medicine, Clinical Research Ethics Committee, Ankara, Turkey.

